# Deep learning reveals prognostic information beyond AJCC stage in routine histopathology of early-stage melanoma: the population-based D-ESMEL study

**DOI:** 10.64898/2026.10.29.26366599

**Authors:** Jasper Ouwerkerk, Farhan Akram, Loes Hollestein, Thamila Kerkour, Catherine Zhou, Andrew P. Stubbs, Antien Mooyaart, Yunlei Li

## Abstract

Of all melanoma patients who develop distant metastasis over time, 41% were initially diagnosed with stage I/II. More accurate prognostic models are needed to select patients for potential (neo)adjuvant therapy. We hypothesise that histopathological features in whole slide images (WSIs) from primary early-stage melanomas provide additional information beyond American Joint Committee on Cancer (AJCC) stage.

We utilised data from the population-based Dutch Early-Stage Melanoma (D-ESMEL) study. This included a discovery set (n=328) of case-control sets matched on follow-up time and known prognostic features (i.e. Breslow thickness, ulceration, age, and sex) and a validation cohort based on >5,500 patients following a nested case-control (NCC) design with stage-matched controls (n=266). We developed an end-to-end deep learning workflow to automatically extract relevant patches from WSIs and predict patient-level metastatic risk based on histopathological features independent of AJCC stage. The discovery set served as a training set for the workflow using 10-fold cross-validation. The prognostic performance of this model and its independence from clinical risk factors was evaluated in the validation cohort using odds ratios (OR) and the weighted C-index.

We achieved a balanced accuracy of 0·88 (µ: 0·81; 95% CI: 0·77–0·85) in the best validation fold of the discovery set. In the validation cohort, metastatic risk predictions from the deep learning model were an independent prognostic factor (adjusted OR: 1·97; *p*<0·01). Combining the deep learning predictions (C-index: 0·86) with AJCC stage (C-index: 0·80) yielded the highest C-index of 0·87, significantly improving the C-index compared to AJCC stage alone (*p*<0·001).

Our histopathology-based deep learning model was rigorously evaluated in a population-based cohort and demonstrated added prognostic value beyond AJCC stage. Further investigations of the underlying biological features could improve clinical decision making and patient outcomes.

## Introduction

Although most early-stage melanoma patients have a favourable prognosis, melanoma-related mortality in this group accounts for 41% of all melanoma deaths.^1,2^ Identifying the minority of stage I/II patients at high risk of metastatic progression remains a major clinical challenge. Recent developments in (neo)adjuvant therapy have increased the clinical importance of refining prognostic assessment. Although adjuvant therapy improves recurrence-free survival in resected stage IIB/IIC/III melanoma, an overall survival benefit has not yet been demonstrated in IIB/IIC patients.^3–5^ Due to the lack of overall survival benefit of adjuvant therapy and the low risk of sentinel lymph node metastasis in T1-T2a melanomas, clinical guidelines in some countries even advise against performing a sentinel lymph node biopsy in this group.^6^ Neoadjuvant therapy showed improved event-free survival compared to adjuvant therapy in stage III melanoma patients and is now applied to stage II in clinical trials based on a biopsy, whereafter immunotherapy is applied before full excision (ClinicalTrials.gov number, NCT05418972).^7,8^ Given these changes in the diagnostic and treatment landscape of melanoma, a test based on the excised primary melanoma is now even more urgently needed to intensify surveillance for low T-stage patients at high risk of metastasis to reduce unnecessary exposure to (neo)adjuvant therapy with severe side-effects among low-risk patients.

Current prognostic assessment of early-stage melanoma is mainly based on Breslow thickness and ulceration, two well-established components of the American Joint Committee on Cancer (AJCC) staging system. Aside from Breslow thickness and ulceration, other factors in early-stage melanoma that might contribute to prognosis include: age, tumour location, WHO subtype, mitosis, regression, lymphovascular invasion, sentinel lymph node status, and Clark level.^9,10^ However, many of the histopathological factors require manual scoring, which may lead to inter-observer variability. This was one of the reasons why mitosis was removed from AJCC staging, despite its prognostic value.^11,12^

As a potential alternative, deep learning approaches can be used to extract data-driven histopathological features from digitised tissue slides, reducing reliance on manual assessment. Several studies have shown that deep learning approaches are promising for prediction of prognosis in melanoma. ^13–17^ However, most studies provided limited insight into the histopathological features underlying model predictions.^13–16^ While one study did characterise these features and showed prognostic value beyond AJCC stage, its reliance on fixed feature representation extracted using a pre-trained model on institutional cohorts may have limited the ability to identify additional histopathological characteristics relevant to prognosis on the population-level.^17^ Histopathological foundation models offer another approach. These large models are pre-trained on H&Es of diverse tissues and can be adapted to multiple prediction tasks across different cancer types. However, previous work suggests that foundation models perform better on diagnostic tasks than on prognosis tasks, for which reported performance remains modest.^18^ Therefore, fixed feature representations extracted using pre-trained models may not fully capture histopathological features relevant to metastatic risk in early-stage melanoma.

We aim to develop a task-specific model in which feature extraction can be optimised directly for metastatic risk prediction.^19^ We utilised histopathological slides from the population-based Dutch Early-Stage Melanoma (D-ESMEL) Study, which was specifically designed for early-stage melanoma prognostic modelling. This study includes a discovery set and a validation cohort with >350 metastatic events in total.^20^ This large dataset provides a unique opportunity to investigate whether a task-specific deep learning model can uncover novel and clinically relevant histopathological patterns associated with metastatic risk in early-stage melanoma patients.

## Material and Methods

### Data

The D-ESMEL study was designed to discover and validate novel prognostic markers for early-stage melanoma and is based on nationwide linked cancer registry and pathology data.^20^ The study included a discovery set where 221 melanoma patients who developed distant metastasis during follow-up (cases) were matched 1:1 to patients who remained distant metastasis-free (controls) on Breslow thickness, ulceration status, age, sex and follow-up time. The validation cohort was based on a population-based cohort of >5,500 stage I/II melanoma patients following a nested case-control (NCC) design including 154 case-control sets with AJCC substage-matched (IA/IB/IIA/IIB/IIC) controls (**figure 1A**). All controls were selected using incidence density sampling, requiring potential controls to be at risk and event-free at the time of the matching case’s event. This design ensures that the observed odds ratios mathematically approximate the incidence rate ratios of the full cohort.^21^ A detailed explanation of the complete design of the D-ESMEL study has been published elsewhere.^20,22^ In this study only haematoxylin and eosin (H&Es) whole slide images (WSIs) of resections were included (i.e., no biopsies) to reduce technical variability. More details on image acquisition are described in **appendix p1**. Afterwards case-control sets were excluded in the discovery set if the control developed distant metastasis after the follow-up period and H&Es were stringently checked for quality and slides with folds, blur, or high levels of melanin covering the tumour area were excluded (**figure 1B**).

**Figure 1.**
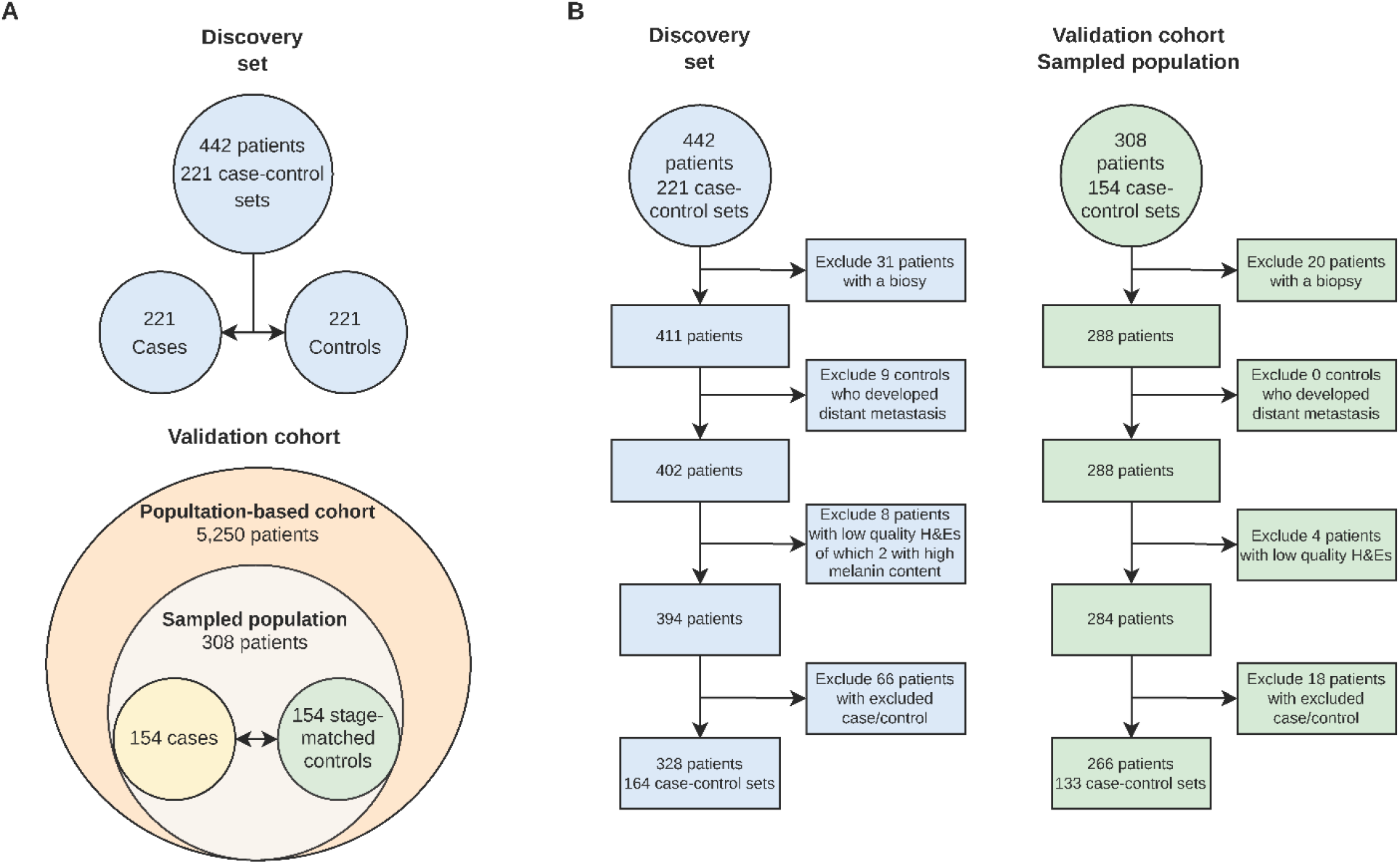
Overview of D-ESMEL study datasets and inclusion criteria. (A) All H&E in the discovery set and the validation cohort selected from the D-ESMEL study before filtering. The validation cohort includes a population-based cohort from which stage-matched case-control sets are sampled. (B) After filtering, the H&Es of the discovery set and validation cohort only include resections of complete case-control sets with high-quality H&Es where controls did not develop metastasis during follow-up. D-ESMEL=Dutch Early-Stage MELanoma. H&E=Haematoxylin and eosin. NCC=Nested case-control.

### Model Development

The H&E WSIs underwent foreground-aware stain normalisation and tumour regions of interest were automatically identified using K-means clustering of 128×128-pixel patches (**figure 2A**). We developed a domain-adversarial multiple instance learning (MIL) framework to generate patient-level representations (*Fp*) predictive for prognosis of distant metastasis (**figure 2B**).^23^ A task-specific feedforward deep learning model was trained on the patient representations (*Fp*) using a stratified 10-fold cross-validation of the discovery set to split the case-control sets into train and validation folds. Afterwards the model was applied to the validation cohort (**figure 2C**). The model generated a risk score ranging from zero to one for each patient. Scores ≤0·50 were classified as low risk of metastasis, and scores >0·50 were classified as a high risk of metastasis. Further details on model development, optimisation, and the software used are provided in the supplementary methods (**appendix p1**).

**Figure 2.**
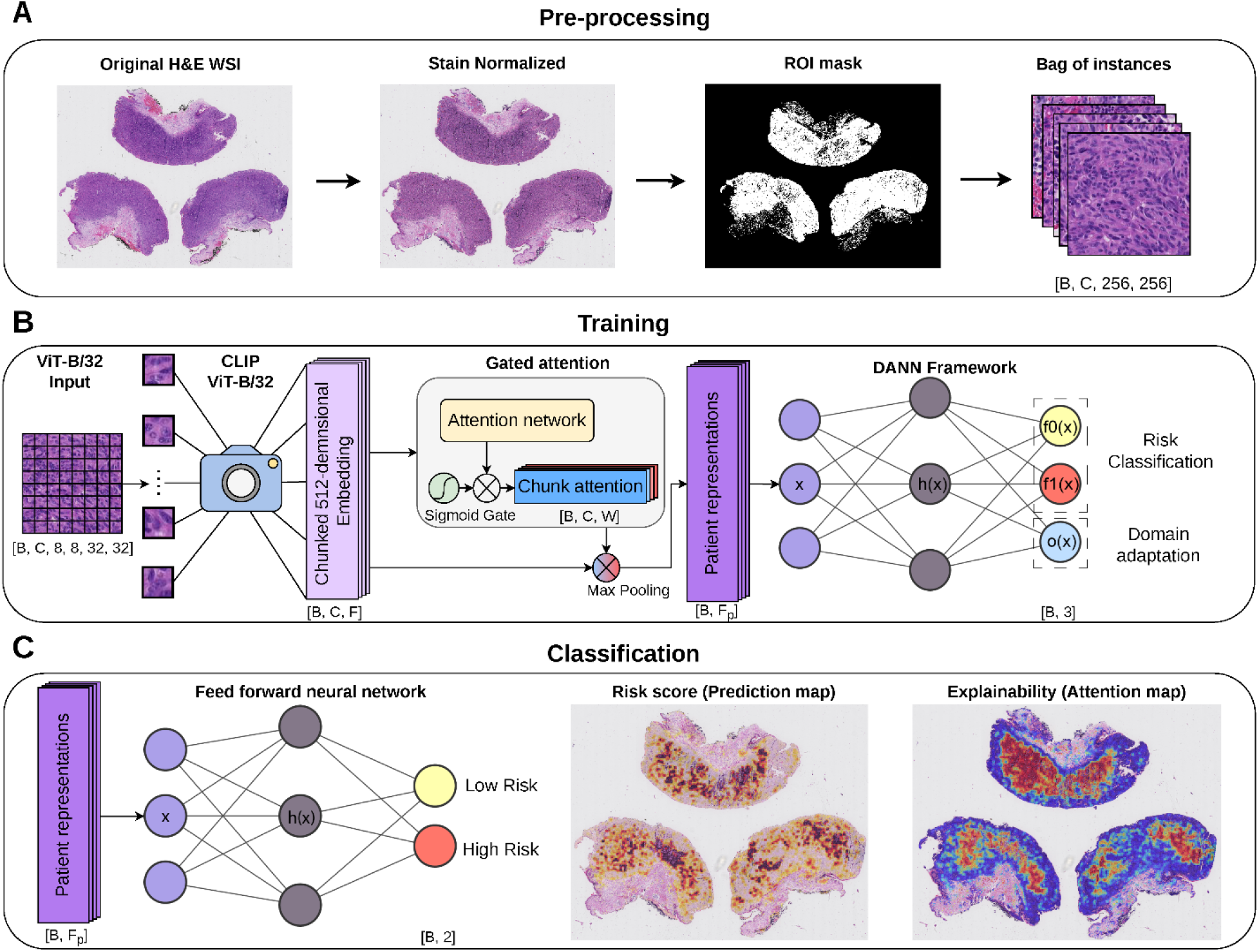
Overview of the end-to-end workflow from pre-processing H&E WSIs to extracting relevant patches and predicting metastatic risk. (A) The H&E WSIs were stain-normalised and relevant patches were automatically extracted and put into bags (*B*) of chunks (*C*). (B) Chunked embeddings (*F*) were generated using a CLIP ViT-B/32 encoder and attention weights (*W*) were learned using a MIL-based approach. Weighted chunks were aggregated using max-pooling to generate patient-level representations (*Fp*). These representations were further refined using a DANN framework. (C) A feed-forward neural network was trained to predict metastasis at the patient-level and to generate attention and prediction heatmaps for explainability. DANN=Domain-Adversarial Neural Network. H&E=Haematoxylin and eosin. MIL=Multiple instance learning. WSI=Whole slide image.

### Statistical Analysis

In the discovery set sensitivity, specificity, balanced accuracy, and the area under the receiver operator characteristic (AUC) score were calculated to evaluate performance. These metrics are further explained in the supplementary methods (**appendix p2)**. In the discovery set, cases and controls were matched on Breslow thickness, ulceration, age and sex. Therefore, any AUC above 0·50 represents an improvement compared to these known prognostic features.

After model development in the discovery set, univariable and multivariable conditional logistic regression (CLR) models were applied to the NCC sets in the validation cohort. Unweighted CLRs were fitted using distant metastasis as the dependent variable to assess the added value of the deep learning model’s predictions to AJCC stage. The deep learning model’s predicted risk score was rescaled to range from zero to ten. Therefore, odds ratios (OR) correspond to 0·1 increase in predicted probability. The independent variables in the multivariable model included the rescaled deep learning model’s predicted risk score, age, sex, Breslow thickness, and ulceration (in addition to the matching on AJCC substage). Breslow and ulceration are included as these strong predictors have prognostic value even within the AJCC substage.

Next, the statistical interaction between AJCC stage and the deep learning model’s predicted risk score was assessed using a CLR to test for equal performance across stages I and II. Additionally, the effect of potential misclassification of stage III disease, due to the absence of a lymph node biopsy for some patients, was assessed by restricting the analysis to case-control sets with a negative sentinel lymph node (SLN) biopsy (SLNB) status and case-control sets with T1a, T1b, and T2a melanomas (n=154). These patients are either not advised to undergo SLNB (T1a) or have an extremely low risk of SLN metastasis (T1b, T2a: <10%).^29^ Finally, the deep learning model was evaluated on the biopsies of the validation cohort, which were excluded in the development of the model and the main analyses in the validation cohort (n=42) to reduce the impact of technical variations.

To assess the added value of the deep learning model’s predictions in the target population, a sample weighted Cox proportional hazards (CoxPH) model was fitted. Sample weights calculation is described in the supplementary methods (**appendix p2**) and in a previous publication.^30^ The C-index of a model that included the AJCC substage was compared to a model with risk predictions of the deep learning model using the log-likelihood test. To validate the deep learning model’s performance within a 5-year follow-up period, a 5-year time-dependent AUC analysis was performed using the log hazard ratios from the CoxPH models. A *p*-value < 0·05 was considered statistically significant. Software versions used are described in the **appendix p2**.

### Interpretability

Activation and class prediction heatmaps (**figure 2C**) were generated to analyse the extracted features from the H&E-stained slides in the validation cohort. The top 25 cases and 25 controls with the highest and lowest predicted probabilities respectively were selected and interpreted by an expert dermatopathologist (A.M.) for evaluation. Representative slides displaying the extracted features were visualised and interpreted.

## Results

In this study we used high-quality H&E WSIs from the D-ESMEL study, excluding any slides with folds, blur, or high levels of melanin covering the tumour area. This resulted in 164 case-control sets (328 WSIs) in the discovery set for model development and for validation 133 case-control sets (266 WSIs), which are nested within a resized population-based cohort of 4,915 AJCC stage I/II patients **(figure 1A)**. Details on the clinical distribution of these datasets are available in **appendix p3**.

The performance of the deep learning model in predicting metastatic risk was evaluated using multiple performance metrics (**figure 3A**), AUC plots (**figure 3B**), and confusion matrices (**figure 3C**). The training and validation folds showed overall strong performance across all folds. The best validation fold performance was observed in fold 1, which achieved a balanced accuracy of 0·88 and demonstrated strong performance across all other metrics. In this fold the deep learning model also translated well to the validation cohort, reaching a balanced accuracy of 0·64 and an AUC of 0·72 (**figure 3A; figure 3B**), which indicates that the model provides predictive value beyond standard AJCC substage. The confusion matrix of fold 1 shows that performance across the training fold, validation fold, and validation cohort is balanced between cases and controls (**figure 3C**).

**Figure 3.**
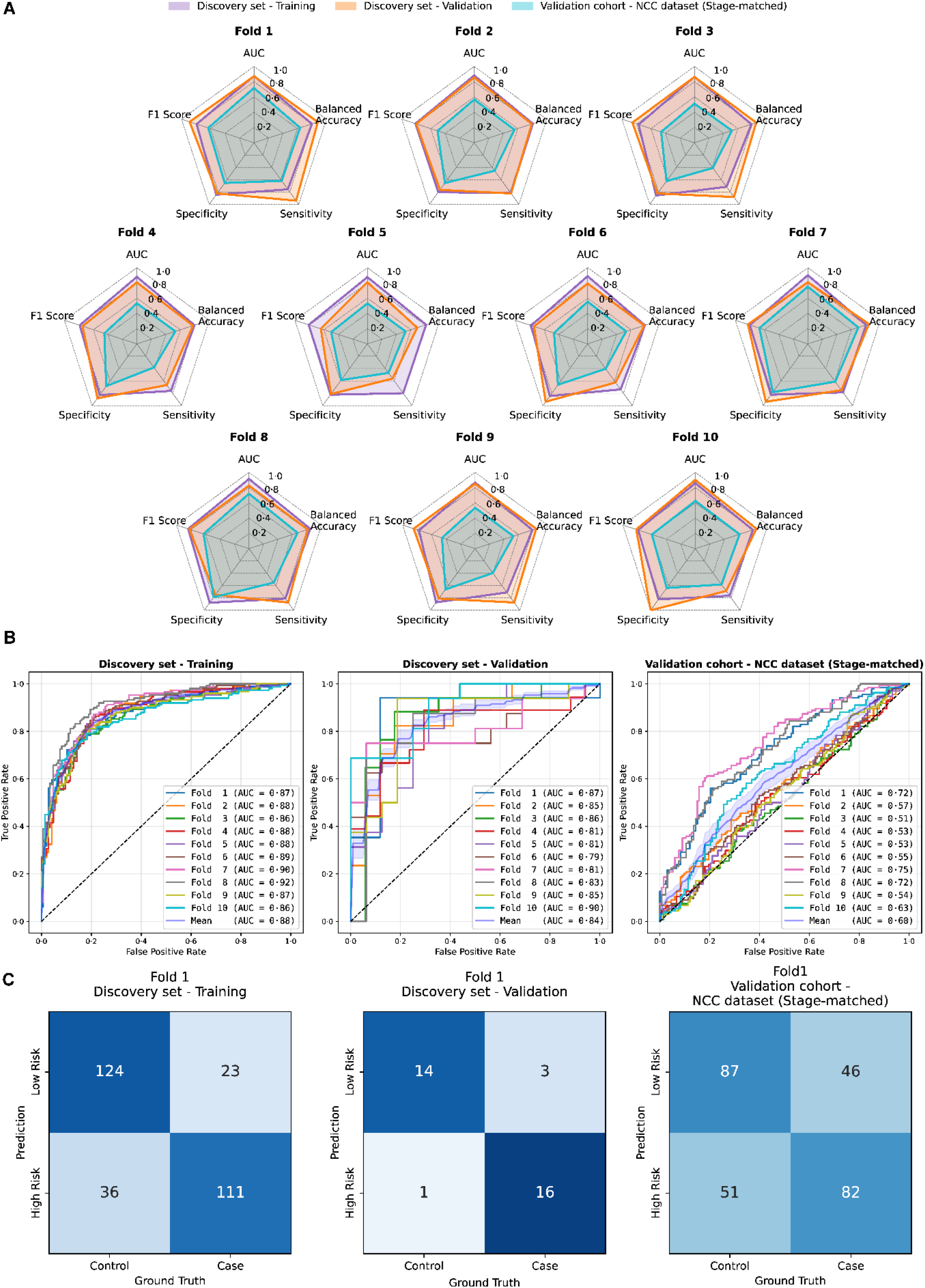
Overview of the deep learning model’s performance across 10 cross-validation folds. (A) The performance across all 10 training/validation folds and the validation cohort. Training/validation fold1 demonstrates the strongest performance based on balanced accuracy. (B) AUC plots reveal strong performance across all 10 folds in the training and validation and an average AUC of 0·60 in the validation cohort. (C) Confusion matrices of the best training/validation fold together with a balanced prediction performance in the validation cohort. AUC=Area under the receiver operating characteristic. NCC=Nested case-control.

An overview of performance of fold 1 model across the AJCC substages in the different datasets is shown in **figure 4**. Based on balanced accuracy relatively equal performance across stages I and II was achieved, but in the validation cohort a low sensitivity (0·29) and high specificity (0·88) were observed in stage I and the opposite was found in stage II (sensitivity: 0·89; specificity: 0·51) (**figure 4A**). The deep learning model tends to predict lower risks for earlier stages and higher risks for later stages even though the effect of stage was largely matched out in the discovery set (**figure 4B**).

**Figure 4.**
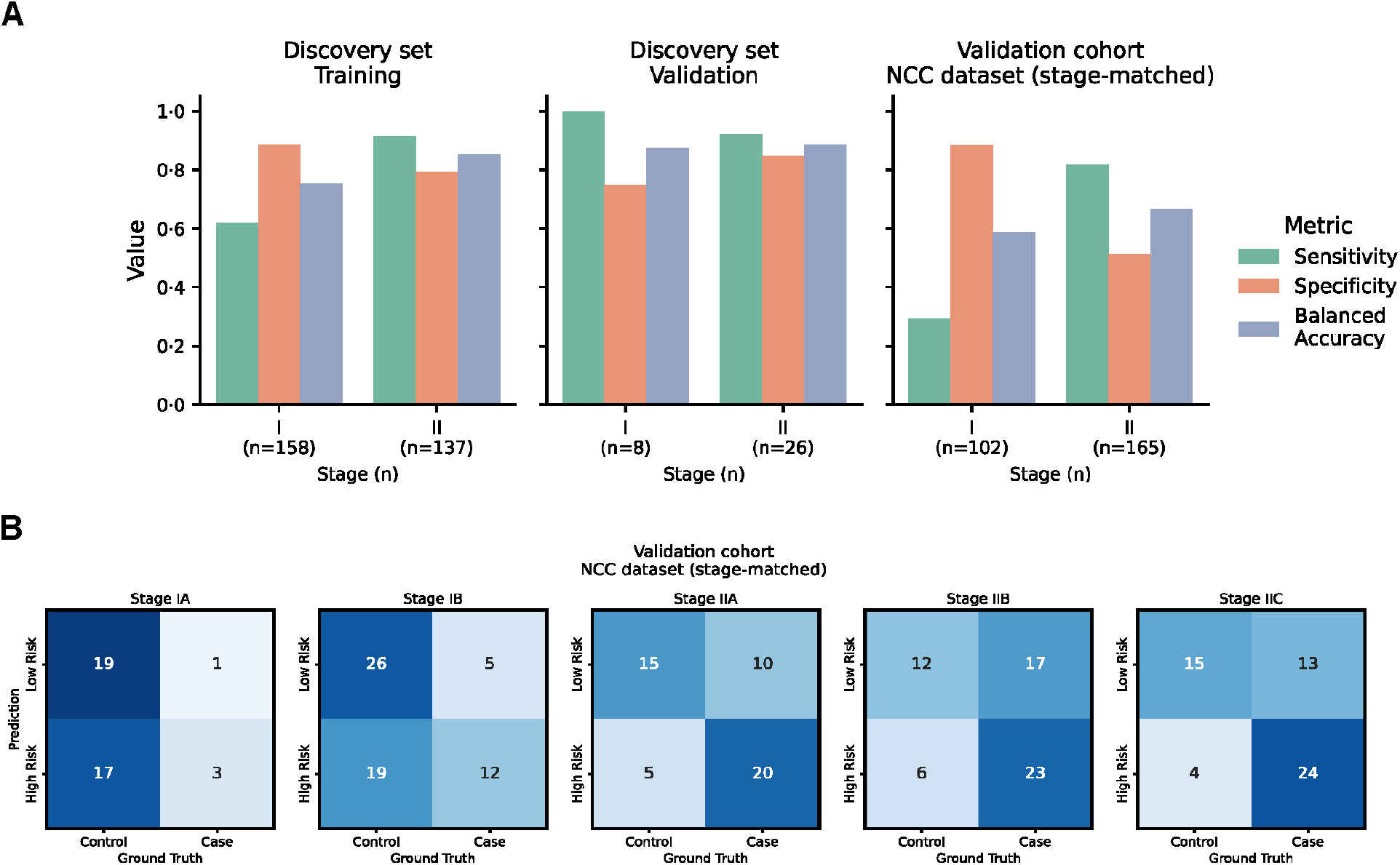
Overview on performance of the deep learning model in fold 1 between cases and controls across stages. (A) Sensitivity, specificity, and balanced accuracy across the training fold, validation fold, and validation cohort. (B) Confusion matrices of each stage in the validation cohort. Generally, the deep learning model predicts lower risk for earlier stages (IA/B) and higher risk for later stages (IIA/B/C). NCC=Nested case-control.

Despite a lower sensitivity in stage I, the deep learning model was significantly associated with increased odds of metastasis in the validation cohort (OR 1·97, 95% CI 1·56-2·50) (**Table 1**). After additional adjustment for Breslow thickness and ulceration, the association remained significant (adjusted OR 1·99, 95% CI 1·56-2·55).

**Table 1.** Odds ratios including the deep learning model’s predicted risk and relevant clinical variables in the validation cohort. CI=Confidence interval. H&E=Haematoxylin and eosin. OR=Odds ratio.

| Characteristics | Univariable |  |  | Multivariable Matched factors |  |  |
| --- | --- | --- | --- | --- | --- | --- |
|  | OR | 95% CI | p-value | OR | 95% CI | p-value |
| *H&E deep learning model* | **1.97** | **1.56 - 2.50** | **<0.01** | **1.99** | **1.56 - 2.55** | **<0.01** |
| *Age* | NA | NA | NA | 1.00 | 0.98 - 1.02 | 0.80 |
| *Sex* | NA | NA | NA | 1.38 | 0.75 - 2.54 | 0.30 |
| *Breslow thickness (mm)* | NA | NA | NA | 1.02 | 0.96 - 1.08 | 0.60 |
| *Ulceration* | NA | NA | NA | 1.61 | 0.59 - 4.37 | 0.35 |

No statistical interaction between AJCC stage and the predicted metastatic risk was found (*p*: 0·21), revealing that the regression coefficient of the deep learning model is similar for both AJCC stage I and II melanoma. This is further supported by the similar balanced accuracies reached between AJCC stage I and II (**figure 4a**). In the subgroup of patients with a negative SLNB or with a very low risk of SLN metastasis (stage T1a/b, T2a), the deep learning model remained prognostic for distant metastasis (adjusted OR: 2·64; 95% CI: 1·68-4·16; *p*: <0·001). In a separate set of biopsies (n=42), of which 27 controls and 15 cases, the deep learning model reached a slightly lower balanced accuracy of 0·59 compared to resections (**figure 3A**). In biopsies, the deep learning model was better at identifying controls than cases, reaching 0·78 specificity, 0·40 sensitivity, and 0·59 AUC (**appendix p4**).

In the sample weighted CoxPH analysis (**figure 5**), the deep learning model showed slightly better prognostic power (C-index: 0·86; CI: 0·80-0·90) for metastatic risk than AJCC stage alone (C-index: 0·80; CI: 0·74-0·85). When the deep learning model was added on top of AJCC stage, a C-index of 0·87 (CI: 0·82-0·91) was reached (**figure 5A**). The added contribution of the deep learning model was statistically significant (*p*: <0·001). The 5-year time-dependent AUC analysis were in alignment with the C-indices where stage reached an AUC of 0·79, the deep learning model an AUC of 0·84, and the combination of the two an AUC of 0·86 (**figure 5B**).

**Figure 5.**
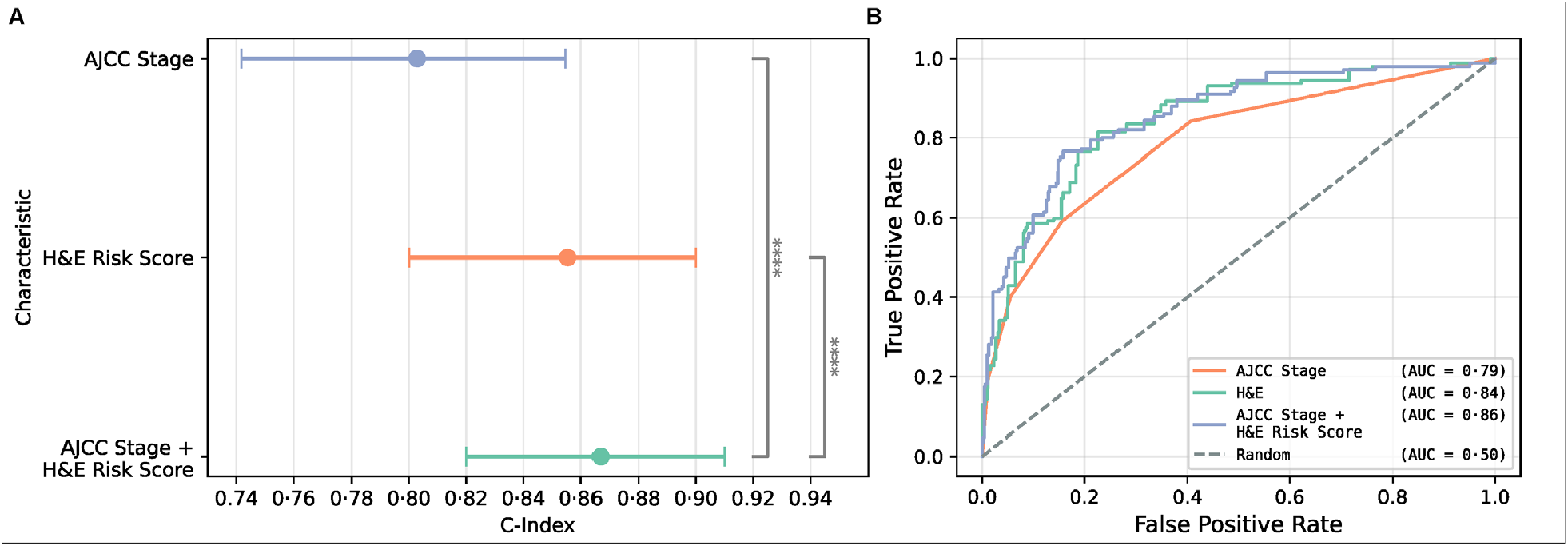
C-indices, standard errors, and AUCs of the deep learning model, AJCC stage, and both variables combined from a weighted CoxPH analysis. (A) The deep learning model significantly adds to AJCC stage achieving a C-index of 0·87. (B) The 5-year time dependent AUC curves demonstrate a similar incremental benefit, reaching an AUC of 0·86 when combining AJCC stage and the deep learning model’s predictions. AJCC=American Joint Committee on Cancer. AUC=Area under the receiver operating characteristic. CoxPH=Cox proportional hazard. H&E=Haematoxylin and eosin.

The interpretation of data-driven histopathological patterns among selected representative controls (**figure 6A**) and cases (**figure 6B**) revealed that attention was highest in the tumour regions. A few patches were also extracted outside the tumour but received very low attention limiting the impact of potential artefacts on the final risk prediction. In the highlighted controls, attention and low-risk predictions co-locate around the tumour border with tumour-infiltrating lymphocytes (TILs) (**figure 6A**). In the highlighted cases, attention and high-risk predictions overlap in ulcerated areas (**figure 6B**), but also contained large regions of low-risk predictions and high attention. Altogether, analysis of the attention heatmaps did not identify a common histopathological pattern that was consistently specific for the selected cases or controls.

**Figure 6.**
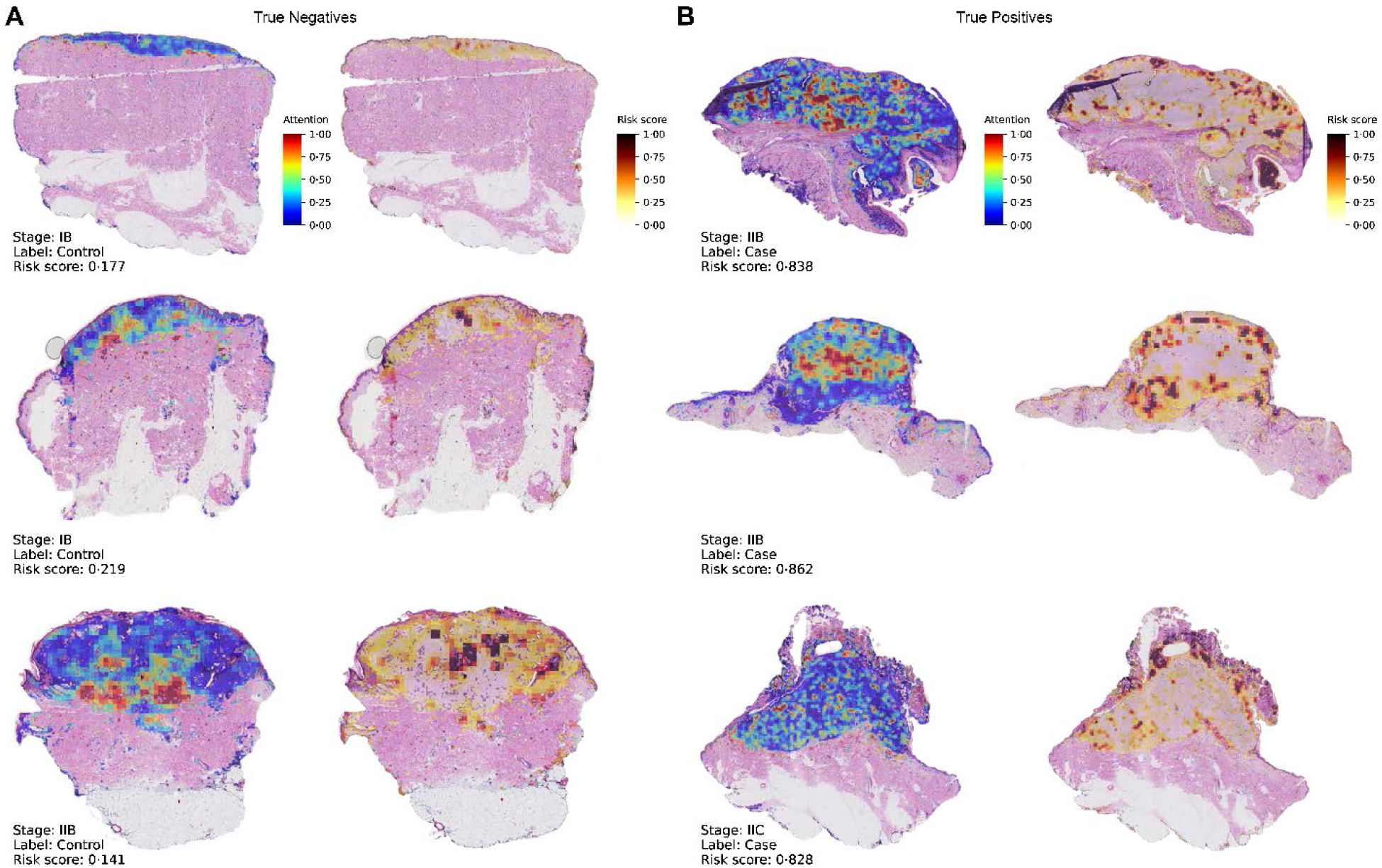
Attention and prediction heatmaps of reconstructed H&E WSIs. (A) Attention and prediction heatmaps across correctly predicted controls (true negatives) generally showing high attention and low risk predictions along the tumour border. (B) Attention and prediction heatmaps across correctly predicted cases (true positives) showing small areas of high attention and high risk predictions around ulcerated areas, but also large regions of low risk predictions with high attention.

## Discussion

We developed and validated an end-to-end workflow based on digital H&E WSI that adds prognostic value for predicting distant metastasis on top of AJCC stage in early-stage melanoma. The workflow was developed on a matched case-control dataset in order to find patterns within the H&E WSIs independent of known prognostic clinical and histopathological factors. Thereafter, we validated the deep learning model predictions in a population-based cohort of >5,500 stage I/II melanoma patients using an efficient NCC design, and demonstrated that the deep learning model added independent and additional prognostic value on top of the AJCC stage. This suggests that critical histopathological features aside from Breslow thickness and ulceration remain underutilised. These features could serve as an alternative or add to manually scored histopathological features and potentially improve prediction of metastasis in early-stage melanoma advancing clinical care and decision making.

Previously, deep learning has been utilised to find data-driven histopathological features to predict disease prognosis of melanomas.^13–17^ Notably, one study also reported prognostic value independent of AJCC stage.^17^ However, these studies were largely based on selected institutional cohorts, making it difficult to establish whether their reported prognostic performance generalises to a nationwide population of patients with early-stage melanoma. In this study, we extended their observations by demonstrating that data-driven histopathological features provide prognostic information beyond AJCC staging in a population-based cohort. This increased the clinical relevance and generalisability of our findings.

The results of our data-driven deep learning approach are consistent with the findings from a biology-driven approach within the D-ESMEL study. In a previous study, we quantified known histological characteristics, such as tumour area, mitosis and TILs. Automated TIL quantification added prognostic value to AJCC staging.^28^ Our findings corroborate those results, as we observed that attention and low-risk predictions co-locate around the tumour border TILs. These observed prognostic patterns were not described in previous studies utilising data-driven histopathological features, possibly due to the use of pre-trained models. These models were pre-trained on either natural images (ImageNet) or on an unknown source of pre-training data and extract fixed feature representations, which may be suboptimal for prognostic prediction.^14–19,31^ Only one of these studies investigated their data-driven features and mainly reported cell morphology patterns, such as higher nuclear pleomorphism and large tumour cell size being associated with adverse survival outcomes, whereas a naevocytoid morphology with smaller cell size and bland monomorphic cells was associated with favourable survival outcomes.^17^ We did not observe these cell morphology patterns to be associated with metastasis in our cohort. However, there was some overlap in the non-cellular morphology reported patterns, as they highlighted patches of fibrosis with dispersed TILs near the tumour border to be associated with lower risk. We also identified patches near the tumour border with TILs in controls. However, fibrosis was not observed consistently enough to be reported as a distinct feature. The lower risk associated with TILs located in peritumoural tissue aligns with another study which found, especially in thin melanomas, that peritumoural CD8+ is associated with better disease free survival.^32^ The overlap with automated TIL quantification plus localisation and the strong standalone performance in an independent population-based validation set, indicated by a C-index of 0·86, suggests that our task-specific model extracts strong histopathological features.

Alongside population-based validation, we examined model performance across each AJCC substage in a matched case-control setting, providing evidence on generalisability on each AJCC substage which was not reported in other studies. We observed only a small difference in overall performance between stage I and II, but did observe a low sensitivity in stage I melanomas (0·29) and low specificity in stage II melanomas (0·51). The difference in performance implies that the model might have learned some stage-correlated morphologies. This highlights the difficulty of predicting rare outcomes, i.e. metastatic events in stage IA melanomas and metastasis-free outcomes in stage IIC melanoma.

Beyond the reported prognostic histopathological patterns in this study, the interpretation and biological understanding of these patterns remain challenging. For instance, the reported patterns were only observed in a subset of samples and do not represent universal prognostic features. Furthermore, some regions lacked an obvious biological pattern both inside and outside the tumour area. However, no strong attention was given to areas with technical artefacts suggesting predictions were based on biological features. Also, we observed large areas of high attention coupled with low-risk predictions in cases. This finding suggests that the attention network may have captured histopathological patterns which resemble those observed in controls, thereby assigning high attention to these regions. Nonetheless, patient-level predictions are formed by the aggregate information contained across the whole tissue rather than by a single histopathological pattern. Therefore, accurate generalisable patient-level risk stratification was still achieved.

Our study has several strengths. Firstly, our end-to-end workflow was developed and evaluated using the D-ESMEL study, which included >350 metastatic events in both a discovery set and an independent validation cohort ensuring robust model development and independent validation. More specifically we utilised a matched case-control design during model development based on Breslow thickness, ulceration, age and sex forcing the deep learning model to learn histopathological features independent of these strong prognostic clinicopathological risk factors. Thereafter a NCC design with stage-matched controls was used to evaluate performance across each AJCC substage and to assess the added prognostic value independent of AJCC stage in a population-based cohort. The validation of our findings in the independent population-based validation cohort further supports that the extracted histopathological signals reflect relevant prognostic information.

Our study also has a few limitations: the low number of biopsies and the use of a single facility for staining and scanning of the H&E WSI limits the generalisability to biopsies and direct application of the model on H&E WSI of other institutes. More biopsies are needed to accurately evaluate the prognostic information in biopsies, which are still clinically relevant as some melanomas are only discovered after assessment of a biopsy by a pathologist whereafter a subsequent resection might contain limited tumour material for analysis. Moreover, additional validation studies based on data from other institutions should establish the robustness of the deep learning model across staining, scanning, and other technical variations.

Our work demonstrated that it is technically feasible to extract additional prognostic information from routinely available H&E-stained slides. This has important clinical implications, as an accurate prognostic test based on the excised primary melanoma could improve risk stratification without requiring additional invasive procedures, such as the SLNB, or costly molecular testing. A major advantage of an H&E-based model is its broad applicability, since digital pathology slides are available for nearly all melanoma patients. Currently, clinicians rely on the SLNB for additional prognostic information beyond T-stage. However, the SLNB is an invasive procedure with possible side effects and a high likelihood of a negative outcome. For this reason, the SLNB will no longer be offered to T1b and T2a melanoma patients in the Netherlands.^29^ This leaves dermatologist reliant solely on only two histological variables (Breslow thickness and ulceration) of T-staging for prognostic information. In other countries, or for patients with higher T-stages, a gene expression profile based on the excised melanoma may be offered to test whether it is safe to forgo SLNB.^33^ While this gene expression profile showed to be relevant to predict a low risk on metastasis, predicting a high risk on metastasis based on gene expression did not outperform AJCC staging.^34^ Our deep learning model adds to AJCC staging, but in a controlled research setting and is therefore not directly applicable in clinical practice. The next step is to translate the data-driven histopathological features into quantifiable histopathological characteristics to transform our findings into actionable prognostic factors, which could potentially assist in selecting patients for increased surveillance or (neo)adjuvant therapy.^3–5,7^ This could be achieved by identifying a combination of hand-crafted histopathological features, defined by expert pathologists, which strongly correlate with the data-driven features. This will involve extensive feature characterisation and validation.

To conclude, we were able to accurately predict prognosis of metastasis in melanoma using histopathological features derived from a task-specific deep learning model. This finding show that routine H&E slides harbour additional prognostic information. Ultimately, translating these findings into clinically actionable prognostic markers could support more informed treatment decisions and improve patient outcomes.

## Data Availability

The clinical data from these patients was provided to the authors by the IKNL. These data are not publicly available and restrictions apply to the availability of the data used for the current study. However, the histopathological images and clinical data are available upon reasonable request to the authors and with permission of the Netherlands Comprehensive Cancer Organisation.

## Author Contributions

Conceptualisation: LH, YL; Data Curation: AM, TK, CZ, LH; Formal analysis: JO, FA; Funding acquisition: AM, YL, LH; Investigation: JO; Methodology: JO, FA, YL; Supervision: LH, AS, FA, YL; Writing – Original Draft: JO, FA; Writing – Reviewing & Editing: LH, FA, AM, AS, TK, CZ, YL.

## Acknowledgements

We would like to thank the Netherland Comprehensive Cancer Organisation (IKNL) for providing the clinical data and the KWF Kankerbestrijding for funding this study.

## Ethics statement

Ethical approval was obtained from the scientific committees of Erasmus MC (MEC-2018–1738), the Dutch Nationwide Pathology Databank, and the Netherlands Cancer Registry.

The use of leftover diagnostic tissue samples for scientific research is based on the ‘no objection’ principle, as outlined in the Code of Conduct for Health Research by the Committee on Regulation of Health Research.

