## Appendix for "Deep learning reveals prognostic information beyond AJCC stage in routine histopathology of early-stage melanoma: the population-based D-ESMEL study"

**Supplementary appendix**

**Supplemental Material: Table of Contents**

### Supplementary Methods

Staining and digitization acquisition

For all patients, formalin-fixed, paraffin-embedded (FFPE) specimens of the primary melanomas of the patients were collected and cut into sections to generate haematoxylin and eosin (H&E) whole slide images (WSIs). The WSIs were scanned at 40x magnification (0·225µm/px) and digitalised with NanoZoomer (Hamamatsu) scanners.

H&E normalization

H&Es were normalized based on a reference colour distribution from four H&E slides of melanomas from the discovery set. Colour statistics (mean and standard deviation) were computed exclusively from foreground pixels in the reference images using intensity thresholding (T=220). The learned transformation was then applied only to the foreground regions in the target WSIs while preserving background characteristics.

Model development

The H&E WSIs underwent foreground-aware stain normalisation to standardise colour distributions. Tumour regions of interest (ROIs) were automatically identified using K-means clustering of 128×128-pixel patches to focus the analysis on diagnostically relevant tissue regions (**Figure 2A**). We developed a domain-adversarial multiple instance learning (MIL) framework to classify primary melanomas as having a high or low risk for metastasis.^1^ Each WSI was divided into non-overlapping 256×256-pixels patches at 10× magnification. These patches were grouped into fixed-size chunks (*C*) of 64 patches and organised into patient-level bags (*B*). Because tumour ROIs varied in size, each patient was represented by a variable number of chunks. Unsupervised patch-level features were learned and extracted using the CLIP ViT-B/32 encoder, which divides each 256×256-pixel patch into an 8×8 grid of non-overlapping image tokens, each corresponding to a 32x32-pixels region. The resulting representations were processed by the Vision Transformer and projected into a 512-dimensional feature embeddings (*F*) for each input patch.^2,3^ These embeddings were then processed using a gated attention mechanism, which assigned importance weights (*W*) to both patches and chunks, to emphasise regions relevant to prognosis.^4^ The weighted chunk features were aggregated using max-pooling to generate a compact patient-level representation (*F_p_*), which was further refined using projection layers and a trainable few-shot adapter to support discrimination between risk groups and adaptation to domain shifts. The resulting representations were passed to Domain-Aware Patient Classifier with two objectives: metastasis classification using cross-entropy loss and domain discrimination via a Gradient Reversal Layer (GRL), following the Domain-Adversarial Neural Network (DANN) framework (**Figure 2B**).^5^ A task-specific feedforward deep learning model was trained on the patient representations using patient-level stratified 10-fold cross-validation within the case-control sets of the discovery sets and subsequently applied to the validation cohort. The model generated a risk score ranging from zero to one for each patient. Scores lower or equal to 0·50 were classified as low risk of metastasis, and scores greater than 0·50 were classified as a high risk of metastasis (**Figure 2C**).

Model optimisation

Training optimized three loss components: (1) classification loss for predictive accuracy, (2) adversarial domain loss to align feature distributions between the discovery set and validation cohort via the GRL, and (3) same-class cross-set alignment loss to cluster samples with identical labels from the discovery set and validation cohort in the latent space. The adversarial weight (λ_adv_) was gradually increased from 0 to 2 to ensure stable training. Separate optimizers (AdamW for classification and Adam/AdamW for the domain branch) were used for the classification and domain branches.

Deep learning models were validated using a stratified 10-fold cross-validation on the case-control sets from the discovery set, where each fold maintained the original clinical matching of case-control sets to prevent data leakage. Model selection was based on Matthews Correlation Coefficient (MCC) with early stopping (patience = 15). The validation cohort remained completely unseen during training and was used only for uniform manifold approximation and projection (UMAP)-based qualitative analysis and final out-of-distribution testing. UMAP features were standardized using z-score normalization prior to evaluation. Reproducibility was ensured through fixed random seeds (42) across PyTorch, CuDNN, NumPy, and Python, with strict verification of no patient overlap.

Cox proportional hazard sample weight calculation

Sample weights were calculated using a generalised linear model (GLM) based on the inverse sampling probability of follow-up time until the last observation. To account for exclusions made in the population-based set, samples within the sampled population were proportionally removed based on the number of included case-control sets in the sampled population to maintain a comparable event rate in this source cohort. Due to the nested case-control design with incidence density sampling, the observed odds ratios mathematically approximate the incidence rate ratios of the full cohort, facilitating analyses of the complete log likelihood tests to assess the added value of the integrated risk score in different multivariable models.

Statistical analysis

Model performance was calculated using the sensitivity, specificity, balanced accuracy, and the area under the receiver operator curve (AUC) score. Sensitivity and specificity represent the accuracy of identifying cases (true positives) and controls (true negatives) respectively. Balanced accuracy is the mean of these two metrics, which shows how balanced prediction performance is between cases and controls. The AUC represents the overall ability of a model to distinguish cases from controls utilising the predicted risk (0·00-1·00).

Software versions

Statistical analyses were performed in R-4.3 using the packages: survival-3.5-8, timeROC-0.4.1, and lmtest-0.9-40.

Supplemental Table 1**:** clinical characteristics of the discovery set and validation cohort. AJJC=American Joint Committee on Cancer. ALM=Acral lentiginous melanoma. DM=Desmoplastic melanoma. LMM=Lentigo maligna melanoma. NM=Nodular melanoma. SSM=Superficial spreading melanoma.

| **Dataset** | **Discovery set** | | **Validation cohort** | |
| --- | --- | --- | --- | --- |
| **Characteristic** | **Case (n=164)** | **Control (n=164)** | **Case (n=133)** | **Control (n=133)** |
| *Time until distant metastasis, years* | | |  |  |
| <2 | 42 (26%) | 0 (0%) | 70 (53%) | 0 (0%) |
| "2-5 | 66 (40%) | 0 (0%) | 60 (45%) | 0 (0%) |
| "5-8 | 42 (26%) | 0 (0%) | 3 (2%) | 0 (0%) |
| >8 | 14 (9%) | 0 (0%) | 0 (0%) | 0 (0%) |
| *Age (median (IQR))* | 63 (50-70) | 62 (51-71) | 68 (56-75) | 69 (55-77) |
| *Sex (n(%))* |  |  |  |  |
| *Female* | 68 (41%) | 68 (41%) | 58 (44%) | 73 (55%) |
| *Male* | 96 (59%) | 96 (59%) | 75 (56%) | 60 (45%) |
| *Breslow, mm (n(%))* |  |  |  |  |
| *≤1·0* | 40 (24%) | 40 (24%) | 20 (15%) | 23 (17%) |
| *>1·0-2·0* | 50 (30%) | 53 (32%) | 33 (25%) | 32 (24%) |
| *>2·0-4·0* | 53 (32%) | 51 (31%) | 33 (25%) | 35 (26%) |
| *>4·0* | 21 (13%) | 20 (12%) | 47 (35%) | 43 (32%) |
| *Ulceration (n(%))* |  |  |  |  |
| *Absent* | 116 (71%) | 116 (71%) | 78 (59%) | 83 (62%) |
| *Present* | 48 (29%) | 48 (29%) | 55 (41%) | 50 (38%) |
| *AJCC Stage (n(%))* |  |  |  |  |
| *IA* | 34 (21%) | 34 (21%) | 20 (15%) | 20 (15%) |
| *IB* | 49 (30%) | 49 (30%) | 31 (23%) | 31 (23%) |
| *IIA* | 34 (21%) | 34 (21%) | 25 (19%) | 25 (19%) |
| *IIB* | 38 (23%) | 38 (23%) | 29 (22%) | 29 (22%) |
| *IIC* | 9 (5%) | 9 (5%) | 28 (21%) | 28 (21%) |
| *Location (n(%))* |  |  |  |  |
| *Face/neck* | 9 (5%) | 8 (5%) | 20 (15%) | 18 (14%) |
| *Lower extremities* | 31 (19%) | 38 (23%) | 33 (25%) | 31 (23%) |
| *Scalp* | 13 (8%) | 7 (4%) | 0 (0%) | 0 (0%) |
| *Trunk* | 87 (53%) | 73 (45%) | 57 (43%) | 51 (38%) |
| *Upper extremities* | 24 (15%) | 38 (23%) | 23 (17%) | 33 (25%) |
| *WHO subtype (n(%))* | |  |  |  |
| *ALM* | 5 (3%) | 7 (4%) | 2 (2%) | 2 (2%) |
| *DM* | 2 (1%) | 0 (0%) | 0 (0%) | 1 (1%) |
| *LMM* | 2 (1%) | 1 (1%) | 3 (2%) | 2 (2%) |
| *NM* | 25 (15%) | 25 (15%) | 29 (22%) | 30 (23%) |
| *SSM* | 129 (79%) | 130 (79%) | 97 (73%) | 91 (68%) |
| *Unspecified* | 1 (1%) | 1 (1%) | 2 (2%) | 7 (5%) |

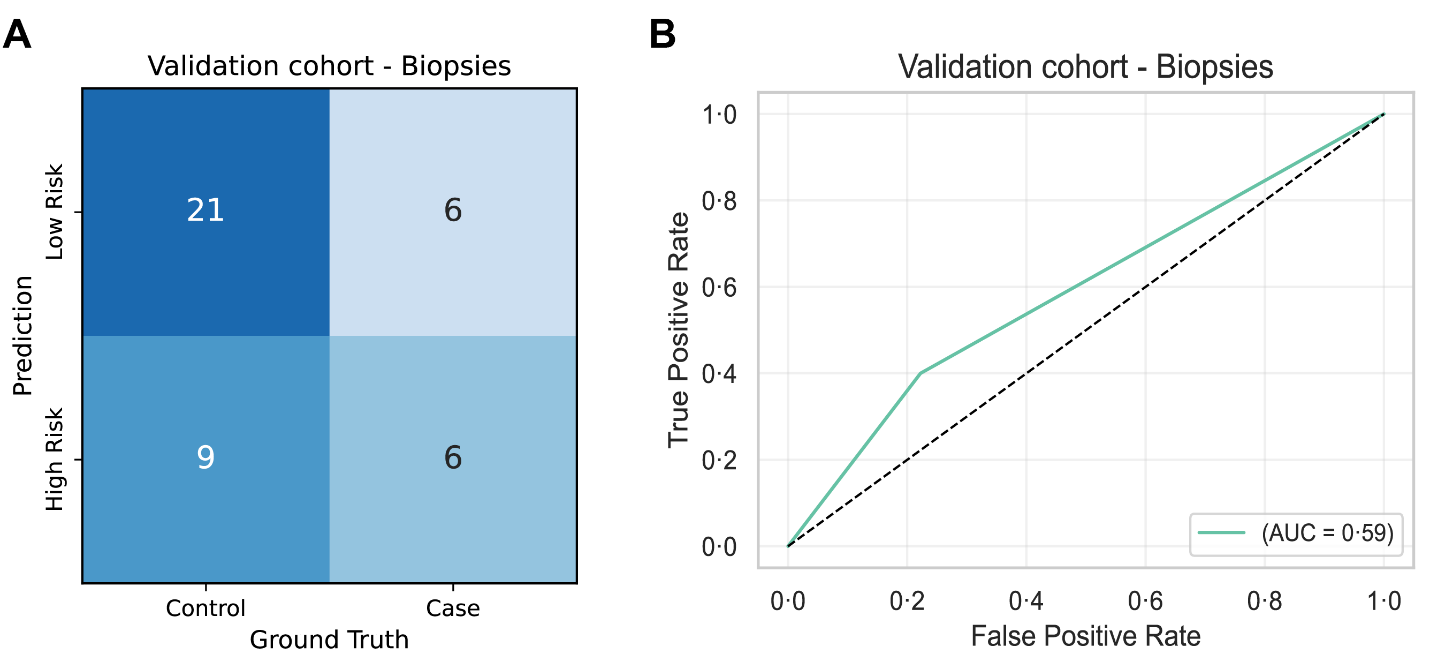

Supplemental Figure 1**: Performance in biopsies of the validation cohort of the deep learning model trained on fold 1**. (A) The confusion matrix reveals controls are easier to predict than cases in biopsies. (B) A relatively low AUC of 0.59 is reached for biopsies. AUC: Area under the receiver operating curve.
